# Genetic Association and Transferability for Urinary Albumin-Creatinine Ratio as a Marker of Kidney Disease in four Sub-Saharan African Populations and non-continental Individuals of African Ancestry

**DOI:** 10.1101/2024.01.17.24301398

**Authors:** Jean-Tristan Brandenburg, Wenlong Carl Chen, Palwende Romuald Boua, Melanie Ann Govender, Godfred Agongo, Lisa K. Micklesfield, Hermann Sorgho, Stephen Tollman, Gershim Asiki, Felistas Mashinya, Scott Hazelhurst, Andrew P Morris, June Fabian, Michèle Ramsay, as members of AWI-Gen and the H3Africa Consortium

**Affiliations:** Sydney Brenner Institute for Molecular Bioscience, Faculty of Health Sciences, University of the Witwatersrand, Johannesburg, South Africa; Strengthening Oncology Services Research Unit, Faculty of Health Sciences, University of the Witwatersrand, Johannesburg, South Africa; National Cancer Registry, National Health Laboratory Service, Johannesburg, South Africa; Clinical Research Unit of Nanoro, Institut de Recherche en Sciences de la Santé, Burkina Faso; Division of Human Genetics, National Health Laboratory Service and School of Pathology, Faculty of Health Sciences, University of the Witwatersrand, Johannesburg, South Africa; Navrongo Health Research Centre, Navrongo, Ghana; Department of Biochemistry and Forensic Sciences, School of Chemical and Biochemical Sciences, C. K. Tedam University of Technology and Applied Sciences, Navrongo, Ghana; SAMRC/Wits Developmental Pathways for Health Research Unit, Faculty of Health Sciences, University of the Witwatersrand, Johannesburg, South Africa; Medical Research Council/Wits University Rural Public Health and Health Transitions Research Unit (Agincourt), School of Public Health, Faculty of Health Sciences, University of the Witwatersrand, Johannesburg, South Africa; African Population and Health Research Center, Nairobi, Kenya; Department of Women’s and Children’s Health, Karolinska Institute, Stockholm, Sweden; Department of Pathology and Medical Sciences, School of Health Care Sciences, Faculty of Health Sciences, University of Limpopo, Polokwane, South Africa; School of Electrical and Information Engineering, University of the Witwatersrand, Johannesburg, South Africa; Centre for Genetics and Genomics Versus Arthritis, Centre for Musculoskeletal Research, The University of Manchester, Manchester, UK; Wits Donald Gordon Medical Centre, School of Clinical Medicine, Faculty of Health Sciences, University of the Witwatersrand, Johannesburg, South Africa

**Keywords:** Chronic kidney disease (CKD), genome-wide association study (GWAS), urinary albumin-creatinine ratio (UACR), African diversity, polygenic risk score

## Abstract

**Background:** Genome-wide association studies (GWAS) have predominantly focused on populations of European and Asian ancestry, limiting our understanding of genetic factors influencing kidney disease in Sub-Saharan African (SSA) populations. This study presents the largest GWAS for urinary albumin-to-creatinine ratio (UACR) in SSA individuals, including 8,970 participants living in different African regions and an additional 9,705 non-resident individuals of African ancestry from the UK Biobank and African American cohorts.

**Methods:** Urine biomarkers and genotype data were obtained from two SSA cohorts (AWI-Gen and ARK), and two non-resident African-ancestry studies (UK Biobank and CKD-Gen Consortium). Association testing and meta-analyses were conducted, with subsequent fine-mapping, conditional analyses, and replication studies. Polygenic scores (PGS) were assessed for transferability across populations.

**Results:** Two genome-wide significant (P<5×10^−8^) UACR-associated loci were identified, one in the *BMP6 region* on chromosome 6, in the meta-analysis of resident African individuals, and another in the *HBB* region on chromosome 11 in the meta-analysis of non-resident SSA individuals, as well as the combined meta-analysis of all studies. Replication of previous significant results confirmed associations in known UACR-associated regions, including *THB53*, *GATM,* and *ARL15*. PGS estimated using previous studies from European ancestry, African ancestry, and multi-ancestry cohorts exhibited limited transferability of PGS across populations, with less than 1% of observed variance explained.

**Conclusion:** This study contributes novel insights into the genetic architecture of kidney disease in SSA populations, emphasizing the need for conducting genetic research in diverse cohorts. The identified loci provide a foundation for future investigations into the genetic susceptibility to chronic kidney disease in underrepresented African populations Additionally, there is a need to develop integrated scores using multi-omics data and risk factors specific to the African context to improve the accuracy of predicting disease outcomes.

## Introduction

Chronic kidney disease (CKD) is a leading risk factor for years of life lost and premature mortality, with a 41.5% relative increase in mortality worldwide from 1990 to 2017 (GBD Chronic Kidney Disease Collaboration et al., 2020; Kovesdy, 2022). The estimated global prevalence of CKD is 9.1% and while predicted to be higher in Sub-Saharan Africa (SSA), the true prevalence and associated risk factors remain understudied (Kaze et al., 2018; GBD Chronic Kidney Disease Collaboration et al., 2020). The Africa Wits-INDEPTH partnership for Genomic Studies (AWI-Gen) cohort, which included ~12,000 participants from four SSA countries in West, East, and Southern Africa, reported overall CKD prevalence as 10.7% (95% confidence interval [CI]: 9.9-11.7), with notable geographic regional differences. The most important risk factors for CKD in SSA were older age, female sex, diabetes, hypertension, and human immunodeficiency virus (HIV) infection (George et al., 2019).

Over the past decade, genome-wide association studies (GWAS) have identified numerous genetic loci associated with kidney function disease, namely, estimated glomerular filtration rate [eGFR], serum creatinine, and urine albumin-creatinine ratio [UACR] (Böger et al., 2011; Pattaro et al., 2012, 2016; Teumer et al., 2016, 2019; Hellwege et al., 2019; Tin and Köttgen, 2020). The majority of the GWAS for kidney function and disease have examined associations with eGFR, while UACR, as a measure for albuminuria, has been investigated less often (Mahajan et al., 2016; Pattaro et al., 2016; Gorski et al., 2017; Haas et al., 2018; Teumer et al., 2019; Wuttke et al., 2019; Zanetti et al., 2019). A recent GWAS in 564,257 individuals of multi-ancestry origins identified 68 associated risk loci for UACR were identified and proposed priority list of genes to explore as targets for the treatment of albuminuria (Teumer et al., 2019). While the majority of kidney disease-associated risk loci have been identified in studies on participants of European and East Asian ancestry, and the African diaspora (Lee et al., 2018), few have focused on participants living in SSA (Böger et al., 2011; Pattaro et al., 2012; Lin et al., 2019; Morris et al., 2019). Recently, in a study of genetic associations of eGFR in a Ugandan population-based cohort, (Fatumo et al., 2020) replicated the association between eGFR and the *GATM* locus.

Replication and transferability of GWAS signals across populations of different ancestries, and specifically with African ancestry populations, tend to be poor despite regional replication often identifying shared associated genomic regions (Pattaro et al., 2012). This may be due to differences in linkage disequilibrium (LD) with the causal variant, allele frequency differences between the populations, underlying population structure, and variabilities in environmental exposures. African populations, with their great genetic diversity and deep evolutionary roots, represent an opportunity for genetic discovery to identify and fine-map disease-associated risk variants (Gomez et al., 2014; Pereira et al., 2021).

Polygenic scores (PGS) are used to quantify and stratify populations according to genetic risk. A PGS based on 63 eGFR-associated alleles showed significant association with kidney disease-related phenotypes, such as chronic kidney failure and hypertensive kidney disease in the Million Veteran Study (US) on 192,868 white and non-Hispanic individuals (Hellwege et al., 2019). A PGS based on 64 urine albumin-to-creatinine ratio (UACR) associated alleles was significantly associated with CKD (Teumer et al., 2019). Further analysis revealed positive associations of the PGS with an increased risk of HT and diabetes. However, PGS often translate poorly across different ancestries (Martin et al., 2017; Kamiza et al., 2022; Kachuri et al., 2023). Since most published GWAS for kidney disease and kidney function markers are based on European ancestry populations, the predictive accuracy of models developed from these studies is expected to be significantly diminished for African populations (Adam et al., 2022; Choudhury et al., 2022; Kamiza et al., 2023; Majara et al., 2023).

In this study, we present a GWAS for UACR conducted within resident Sub-Saharan African individuals. This population cross-sectional study includes a cohort of 8,970 individuals from four SSA countries from the AWI-Gen study (Ali et al., 2018), the African Research on Kidney Disease (ARK) study (Kalyesubula et al., 2020), with 9,705 individuals of African-ancestry from the UK Biobank (UKB) and African American participants from the CKD-Gen Consortium (Teumer et al., 2019). The primary objectives are to: (1) identify genetic loci associated with UACR as a marker of kidney disease in individuals from SSA and of African ancestry; (2) explore the replication of findings identified in previous GWAS; (3) perform analysis and comparison of PGS derived from non-African and multi-ancestry population studies and evaluate their transferability to African populations.

## Participants and Methods

### Study participants

#### Africa Wits-INDEPTH Partnership for Genomic Research (AWI-Gen)

The study participants are a subset of the population cross-sectional AWI-Gen study (Ramsay et al., 2016; Ali et al., 2018). The study recruited adults primarily between the ages of 40 and 80 years from six SSA study sites in West Africa (Nanoro, Burkina Faso and Navrongo,Ghana), East Africa (Nairobi, Kenya) and in South Africa (Bushbuckridge - hereinafter referred to as Agincourt Mpumalanga Province, Dikgale, Limpopo Province; and Soweto, Gauteng Province). All participants were of self-identified black ethnicity. Data collection was described in detail previously (Ali et al., 2018; George et al., 2019). Detailed demographic data, health-related questionnaire data, and anthropometric measurements were collected. Peripheral blood samples and urine samples were collected for biomarker assays (the relevant assays are described below). DNA was extracted from peripheral blood-derived buffy coat samples and used for genotyping. Urine microalbumin was measured using a colorimetric method on the Cobas© 6000/c501 analyzer, and urine creatinine was measured by the modified Jaffe method(Craik et al., 2023). This study was approved by the Human Research Ethics Committee (Medical), University of the Witwatersrand, South Africa (M121029, M170880) and the ethics committees of all participating institutions. All participants provided written informed consent following community engagement and individual consenting processes.

#### African Research on Kidney Disease (ARK)

The African Research Kidney Disease (ARK) study is a well characterised population-based cohort study of 2021 adults (20–80 years) of self-identified black ethnicity from Agincourt, (Mpumalanga, South Africa) with demographic data, health-related questionnaire data, and anthropometric measurements collected at enrolment (Fabian et al., 2022). Blood and urine were collected for biomarker assays (the relevant assays are described below). DNA was extracted from buffy coat samples and used for genotyping. Urine microalbumin was measured using a colorimetric method on the Cobas© 6000/c501 analyzer, and urine creatinine was measured by the modified Jaffe method(Craik et al., 2023). This study was approved by the Human Research Ethics Committee (Medical), University of the Witwatersrand, South Africa (M160939). All participants provided written informed consent following community engagement and individual consenting processes. The geographical area of recruitment overlaps with the Agincourt sub-cohort of AWI-Gen but there is no overlap in participants.

#### UK Biobank (UKB)

Individuals of self-reported Caribbean and African ancestry from the UKB were identified for this study. Of this subset of UKB individuals, those with both genotyping and UACR data were retained for the analysis. UACR was derived using urinary levels of albumin and creatinine. In the UKB, albumin was measured using the immuno-turbidimetric analysis method (Randox Biosciences, UK) while creatinine was measured using the enzymatic analysis method (Beckman Coulter, UK) (Casanova et al., 2019).

### Phenotype generation and harmonization

UACR was calculated for AWI-Gen and ARK studies using urinary levels of albumin and creatinine as previously described (George et al., 2019; Fabian et al., 2022). Participants with missing values for albumin and creatinine were excluded from this study. We applied filtering criteria similar to those employed by the CKD-Gen consortium (Köttgen and Pattaro, 2020). In cases where the values for urine albumin and urine creatinine fell outside the upper and lower limits of detection, the values were replaced with the respective upper and lower limits: for urine creatinine, the range was 3 to 400 mmol/L and for urine albumin, the range was 3.75 to 475 mg/L for AWI-Gen and ARK. For the UKB dataset, the upper limit was 6.7 mg/L for urine albumin. Albuminuria was defined as UACR >3.0mg/mmol.

### Genotyping

#### AWI-Gen and ARK

Genomic DNA was genotyped using the H3Africa custom genotyping array. The H3Africa custom array was designed as an African-common-variant-enriched GWAS array (https://www.h3abionet.org/h3africa-chip) (Illumina) with ~2.3 million single nucleotide polymorphisms (SNPs).

#### UK-Biobank

Genotyping was performed by Affymetrix on two closely related purpose-designed arrays. ~50,000 participants were genotyped using the UK BiLEVE Axiom array (Resource 149600) and the remaining ~450,000 were genotyped using the UK Biobank Axiom array (Resource 149601). The dataset is a combination of results from both arrays. A total of 805,426 markers were released in the genotype data. We extracted individuals with self-reported (Data-Field in dataset 21000) African Ancestry split between African (UKB-African) and Caribbean origins (UKB-Caribbean) from the raw dataset (Casanova et al., 2019).

#### Quality control

For each dataset, AWI-Gen (Choudhury et al., 2022), ARK and UKB, the following pre-imputation quality control (QC) steps were applied: removal of non-autosomal and mitochondrial SNPs; SNPs with genotype missingness greater than 0.05; minor allele frequency (MAF) less than 0.01; and Hardy-Weinberg equilibrium (HWE) P-value less than 0.0001. Individuals were excluded if they had more than 5% overall genotype missingness; heterozygosity lower than 0.150 and higher than 0.343; and discordant genotype and phenotype sex information. We used the GWAS QC workflow of the H3Africa Consortium Pan-African Bioinformatics Network to perform data QC (H3ABioNet H3AGWAS) (https://github.com/h3abionet/h3agwas) (Baichoo et al., 2018; Brandenburg et al., 2022a).

In our final QC step, we identified and excluded outliers, admixed and related individuals using PCASmart, a feature of the EIGENSOFT software (Price et al., 2006), Admixture software (Alexander et al., 2009) using AGV (Gurdasani et al., 2015) and 1000 Genomes Project data (Auton et al., 2015) and PLINK (Version 1.9) (Purcell et al., 2007; Chang et al., 2015). More detail on filter parameters for each software can be found in **Supplementary Table 1**.

#### Imputation

Genotype imputation was performed on each dataset separately (AWI-Gen, ARK, and UKB) using the Sanger Imputation Server with the African Genome Resources reference panel (https://www.sanger.ac.uk/tool/sanger-imputation-service/). EAGLE2 was used for pre-phasing and the PBWT algorithm was used for imputation (Loh et al., 2016). After imputation, poorly imputed SNPs with info scores less than 0.3 and with a HWE P-value less than 1×10^−04^ were removed. The genomic positions were mapped to GRCh37p11.

#### Phenotype transformation for association testing

For AWI-Gen, ARK, and UKB datasets, UACR was transformed on the logarithm scale. Linear regression of variables was performed with covariates in R (Version 3.6): ln(UACR) ~age + sex + genetic principal components (PCs) 1-5. Residuals were extracted and transformed using Rank-Based Inverse Normal Transformation to ensure the normal distribution of residuals (Casanova et al., 2019). PCs were calculated using a sub-set of LD pruned pre-imputed SNPs in PLINK (Version 1.9) (Purcell et al., 2007; Chang et al., 2015). The sub-set was derived by LD pruning using PLINK (Version 1.9) (Purcell et al., 2007; Chang et al., 2015) with an LD (r^2^) threshold of 0.2 with windows of 50 kb and 10 kb for step size.

#### Association testing

Mixed model association testing was performed with imputed genotype probabilities using GEMMA (Version 0.98.1) (Zhou and Stephens, 2012). GEMMA uses a relatedness matrix to account for genetic structure and relatedness between individuals. The relatedness matrix was built with a sub-set of pre-imputed SNPs described above.

Mixed model association testing was performed independently on each dataset. A total of nine datasets were tested. The datasets were defined as follows: six datasets for AWI-Gen: AWI-Agincourt, AWI-Dikgale, AWI-Nanoro, AWI-Nairobi, AWI-Navrongo and AWI-Soweto; one dataset for ARK: ARK-Agincourt; and two datasets for UK Biobank: UKB-Caribbean and UKB-African. For each dataset, Quantile-to-quantile plots (QQ-plots) were generated, and inflation factors were calculated using SNPs with MAF>0.01 to verify that the association signals were not inflated due to unaccounted population sub-structure. The genome-wide significance level for novel discovery was considered at P<5×10^−08^.

#### CKD-Gen

We used previously published meta-analysis summary statistics from the CKD-Gen Consortium. The CKD-Gen Consortium datasets consist of three meta-analysis summary statistics: 1) CKD-Gen European ancestry individuals (CKD-Gen-EA); 2) CKD-Gen African American ancestry individuals (CKD-Gen-AA); and 3) CKD-Gen Multi-ancestry individuals (CKD-Gen-MA) which include individuals from CKD-Gen-EA and CKD-Gen-AA (Teumer et al., 2019). The CKD-Gen Consortium meta-analysis summary statistics were retrieved from http://ckdgen.imbi.uni-freiburg.de/.

Briefly, CKD-Gen-AA is a meta-analysis based on 7 studies with African American participants. For each study, genotyping was performed using genome-wide arrays followed by application of study-specific quality filters prior to phasing, imputation, and association analysis software (description can be found in **Supplementary Table 1 and 2** from (Teumer et al., 2019)). Meta-analysis was performed using fixed effects inverse-variance weighted meta-analysis of the study-specific GWAS result files with imputation quality (IQ) score > 0.6 and MAC > 10, effective sample size ≥ 100, and a beta < 10, using METAL (for more details see (Teumer et al., 2019)).

### Meta-analysis

Fixed-effect meta-analyses were conducted using the METASOFT software (Han and Eskin, 2011). The first meta-analysis (Meta_SSA_) used the GWAS summary statistics generated from individual-level data from resident SSA populations. This included AWI-Agincourt, AWI-Dikgale, AWI-Nanoro, AWI-Nairobi, AWI-Navrongo, AWI-Soweto and ARK-Agincourt. The second meta-analysis (Meta_NONRES_) included data from individuals of African ancestry who are not residing in SSA. We used the GWAS summary statistics generated from individual-level data from the UK-Biobank (UKB-African and UKB-Caribbean) and CKD-Gen African American sub-set (CKD-Gen-AA). The third meta-analysis (Meta_ALL_) consisted of a meta-analysis that pooled the summary statistics of all studies from AWI-Agincourt, AWI-Dikgale, AWI-Nanoro, AWI-Nairobi, AWI-Navrongo, AWI-Soweto, ARK-Agincourt, UKB-African, UKB-Caribbean and CKD-Gen-AA. **Figure 1** outlines the meta-analysis workflow. As a secondary analysis, role of heterogeneity had been investigated between cohorts from different regions of origin by performing separate meta-analyses for residents of Southern African (AWI-Agincourt, AWI-Dikgale, AWI-Soweto, and ARK-Agincourt) and residents of West Africa (AWI-Nanoro, AWI-Navrongo). Random-effects model from METASOFT (Han and Eskin, 2011) take account for potential heterogeneity between study, we performed Meta RE using all dataset (Meta_ALL_^RE^) (Borenstein et al., 2010; Adriani Nikolakopoulou et al., 2014). The genome-wide significance level for novel discovery was considered at P<5×10^−08^.

**Figure 1:**
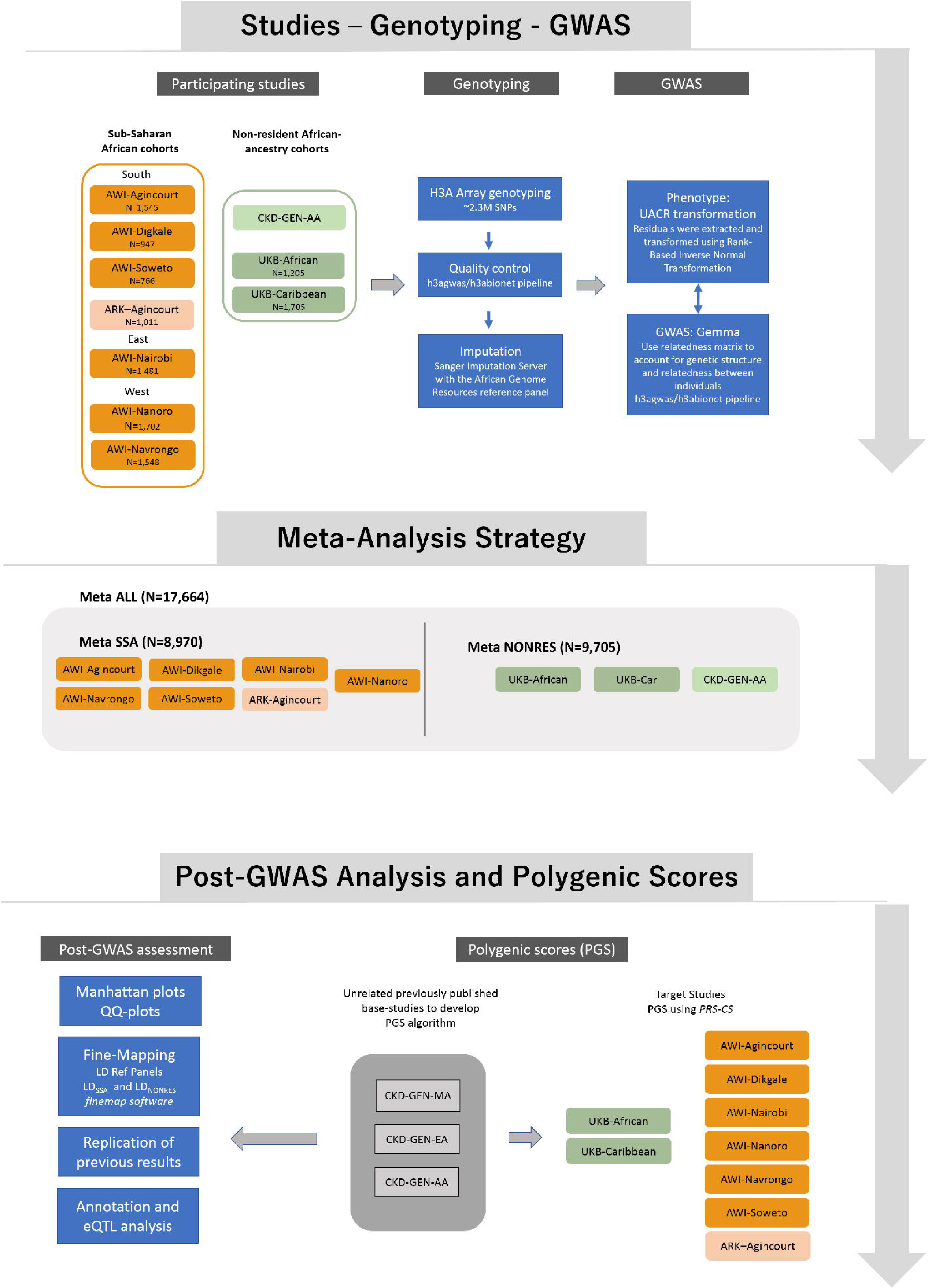
Study design showing data sources, the analysis strategy and post-GWAS analysis approach.

### Post association analysis

#### Plotting

QQ-plots and Manhattan plots were generated using the FastMan library (Paria et al., 2022) (available at https://github.com/kaustubhad/fastman) and the Hudson library (available at https://github.com/anastasia-lucas/hudson). These visualizations were created using SNPs with a MAF threshold of 0.01 or more. For regional plots, we utilized the standalone version of the LocusZoom software (Pruim et al., 2010).

#### Genetic LD reference

For the estimation of the LD reference panel for conditional and joint (COJO) analysis, clumping, and fine-mapping, three LD reference panels were constructed using genotype data from the appropriate datasets. For resident SSA dataset comparisons, the LD reference panel (LD_SSA_) was constructed using AWI-Gen and ARK individual-level genotype data. For non-resident SSA dataset comparisons, the LD reference panel (LD_NONRES_) was constructed using UKB individual-level genotype data. For the combined datasets comparison, the LD reference panel (LD_ALL_) was constructed using AWI-Gen, ARK, and UKB individual-level genotype data.

#### Fine-mapping and lead SNPs

For each locus with a lead SNP with a P-value below 5×10^−08^, fine-mapping was conducted using the H3ABioNet H3AGWAS pipeline and implementing a stepwise model selection procedure through GCTA (Yang et al., 2011, 2012; Brandenburg et al., 2022a) to identify independently associated SNPs. Subsequently, we utilized the FINEMAP software (Version 1.4) (Benner et al., 2016), considering one causal variant, to define the credible set with 99% confidence using a stochastic approach (Benner et al., 2016).

#### Conditional analyses (GCTA)

Conditional analyses used the GCTA software implemented within the H3AGWAS pipeline, with summary statistics obtained from the meta-analyses as input. In these analyses, the lead SNPs identified in each meta-analysis were conditioned upon lead SNPs found in previously published studies. Changes in the p-value, both increasing or decreasing significance, of the lead SNP, confirmed a relationship between the two SNPs.

#### Replication of previous findings

Replication was performed according to the following criteria: 1) Exact replication: if any genome-wide significant lead SNPs found in CKD-Gen-EA and CKD-Gen-MA reached statistical significance (p<0.05) in Meta_SSA_, Meta_NONRES_ or Meta_ALL_ after Bonferroni correction (A total of 60 independent lead SNPs were identified in the CKD-Gen datasets, of which 55 lead SNPs were from CKD-Gen-EA and 57 lead SNPs were from CKD-Gen-MA) and that the lead SNPs have same direction of effect. 2) LD Window replication: for a given genome-wide significant SNP found in the CKD-Gen datasets, SNPs were extracted from Meta_SSA_, Meta_NONRES_ and Meta_ALL_ that are in LD with the said CKD-Gen lead SNP. LD pruning used the clump procedure in PLINK (Version 1.9) (r2 = 0.1, windows size 1000 kb, P_1_ = 5×10^−08^, P_2_ = 0.1). The lowest p-value(s) from SNPs within the given LD window were extracted and this LD window was considered statistically significant if the p-value was less than 5×10^−04^ in both datasets. Additionally, the direction of effect between the CKD-Gen and Meta-datasets (Meta_SSA_, Meta_NONRES_ and Meta_ALL_) must be consistent. Conditional analyses were performed between the genome-wide significant SNP(s) in CKD-Gen and lead SNP in our meta-analyses to confirm the replication.

For replication, the findings from Meta_SSA_ were compared to CKD-Gen-MA and CKD-Gen-EA, and the findings from Meta_NONRES_ and Meta_ALL_ were only compared to CKD-Gen-EA to avoid sample overlaps within the CKD-Gen datasets (as CKD-Gen-AA is contained within CKD-Gen-MA).

#### Annotation and Expression Quantitative Trait Locus (eQTL) analysis

Functional annotation of genome-wide significant SNPs found in Meta_SSA_, Meta_NONRES_ and/or Meta_ALL_ was done using the ANNOVAR software (Wang et al., 2010). eQTL analysis was performed using the database of cis-eQTLs in both glomerular and tubulointerstitial tissues, derived from participants in the Nephrotic Syndrome Study Network (NEPTUNE) using SNPs with false discovery rate (FDR) < 0.05 (Han et al. 2023). In this analysis a 1000kb window was defined around each genome-wide significant locus and an eQTL was considered significant if the LD (r2) was ≥0.01 between the lead SNP and significant eQTL, LD computation used the genetics data from the African populations from the 1000 Genomes Project (v5a, hg19) (Auton et al., 2015; Sudmant et al., 2015).

#### Polygenic Scores

PGS were computed for each dataset independently (AWI-Agincourt, AWI-Dikgale, AWI-Nanoro, AWI-Nairobi, AWI-Navrongo, AWI-Soweto ARK-Agincourt, UKB-African and UKB-Caribbean). The effect sizes from 3 previous studies were used: CKD-Gen-AA, CKD-Gen-MA, and CKD-Gen-EA. PRS-CS (Ge et al., 2019), software that estimates posterior SNP effect sizes by implementing continuous shrinkage (CS) priors, was used to calculate the PGS. As external LD references are required for this analysis, the African LD data derived from the 1000 Genomes Project by the PRScs project was used for this purpose (accessible at https://github.com/getian107/PRScs). The PGS values were regressed against the residualized UACR value in a linear regression model that adjusted for age, sex, and the first five principal components to assess the performance of PGS.

## Results

### Study participants and phenotype data

Genomic and phenotypic data were accessible for 7,959 individuals in the AWI-Gen datasets, 1,011 individuals in the ARK dataset, and 2,916 individuals in the UK-Biobank dataset with 1,205 individuals and 1,711 individuals in UKB-African and UKB-Caribbean respectively (**Supplementary Figure S1**). CKD-Gen AA was a meta-analysis of 7 studies including 6,795 individuals in total. Overall, there was a higher prevalence of albuminuria (17.9%; median UACR 1.01mg/mmol) among individuals from the UKB with African and Caribbean ancestry compared to individuals residing in SSA, where notable regional differences were observed. The highest prevalence of albuminuria occurred in AWI-Agincourt, South Africa (14.1%; median UACR 0.59mg/mmol) while the lowest prevalence occurred in AWI-Nanoro, West Africa (prevalence of albuminuria 4.5%, median UACR 0.35 mg/mmol) (**Table 1**).

**Table 1:** Study participants and phenotype data. Participant characteristics for each AWI-Gen study site, ARK-Agincourt and UKB-African and UKB-Caribbean, with phenotype distributions of UACR (median) and covariables used in the study.

| Dataset | Sample size | Country of residence | <sup>1</sup>Age (years) | Males (%) | <sup>2</sup>UACR | <sup>3</sup>Albuminuria (%) |
| --- | --- | --- | --- | --- | --- | --- |
| **Non-resident in <sup>4</sup>SSA** | **9,705** |  |  |  |  |  |
| <sup>5</sup>CKD-Gen-AA | 6,795 | USA |  |  |  |  |
| <sup>6</sup>UKB-African | 1,205 | UK | 51.8 | 48.5 | 1.10 (1.70) | 20.0 |
| <sup>7</sup>UKB-Caribbean | 1,711 | UK | 52.8 | 59.6 | 0.93 (1.44) | 16.4 |
| <sup>8</sup>UKB-All | 2,916 | UK | 52.4 | 55.0 | 1.01 (1.55) | 17.9 |
| **Resident in SSA** | **8,970** |  |  |  |  |  |
| <sup>9</sup>AWI-Nanoro | 1,702 | Burkina Faso | 49.6 | 50.8 | 0.35 (0.44) | 4.9 |
| <sup>10</sup>AWI-Navrongo | 1,548 | Ghana | 51.1 | 54.4 | 0.41 (0.48) | 6.7 |
| <sup>11</sup>AWI-Nairobi | 1,481 | Kenya | 48.8 | 54.0 | 0.63 (0.67) | 11.4 |
| <sup>12</sup>AWI-Agincourt | 1,545 | South Africa | 54.3 | 59.8 | 0.59 (0.99) | 14.1 |
| <sup>13</sup>AWI-Dikgale | 917 | South Africa | 52.1 | 68.3 | 0.63 (0.51) | 10.4 |
| <sup>14</sup>AWI-Soweto | 766 | South Africa | 49.5 | 100.0 | 0.36 (0.57) | 11.4 |
| AWI-All | 7,959 |  | 50.9 | 51.0 | 0.48 (0.51) | 9.5 |
| <sup>15</sup>ARK-Agincourt | 1,011 | South Africa | 38.8 | 58.0 | 0.57 (0.94) | 13.4 |
| **Total** | **17,664** |  | **50.3** | **52.6** | **0.59 (0.85)** | **11.9** |

<sup>1</sup>data reported as the mean.
<sup>2</sup>UACR: urine albumin: creatinine ratio: mg/mmol; reported as median (interquartile range).
<sup>3</sup>Albuminuria: UACR>3.0mg/mmol.
<sup>4</sup>SSA: Sub-Saharan Africa.
<sup>5</sup>CKD-Gen Consortium: African American Ancestry individuals. CKD-GEN-AA data are summary statistics of meta-analysis from Teumer et al. downloadable at the CKD-GEN consortium website (http://ckdgen.imbi.uni-freiburg.de/), information relative to samples had been reported in relevant papers.
<sup>6</sup>UKB Biobank: Individuals of self-reported African Ancestry; <sup>7</sup>self-reported Caribbean Ancestry; <sup>8</sup>self-reported African and Caribbean Ancestry.
<sup>9</sup>Africa Wits -INDEPTH partnership for Genomic Studies (AWI): individuals of African Ancestry from <sup>9,10</sup>West Africa; <sup>11</sup>East Africa; <sup>12</sup>-South Africa.
<sup>15</sup>African Research on Kidney Disease (ARK) Study: South Africa

### Meta-analysis

Meta-analyses were conducted to investigate the genetics of UACR in resident Sub-Saharan African datasets (Meta_SSA_) (**Figure 2a**), non-resident Sub-Saharan African datasets (Meta_NONRES_) (**Figure 2b**) and all African ancestry datasets (Meta_ALL_) (**Figure 2c**).

**Figure 2:**
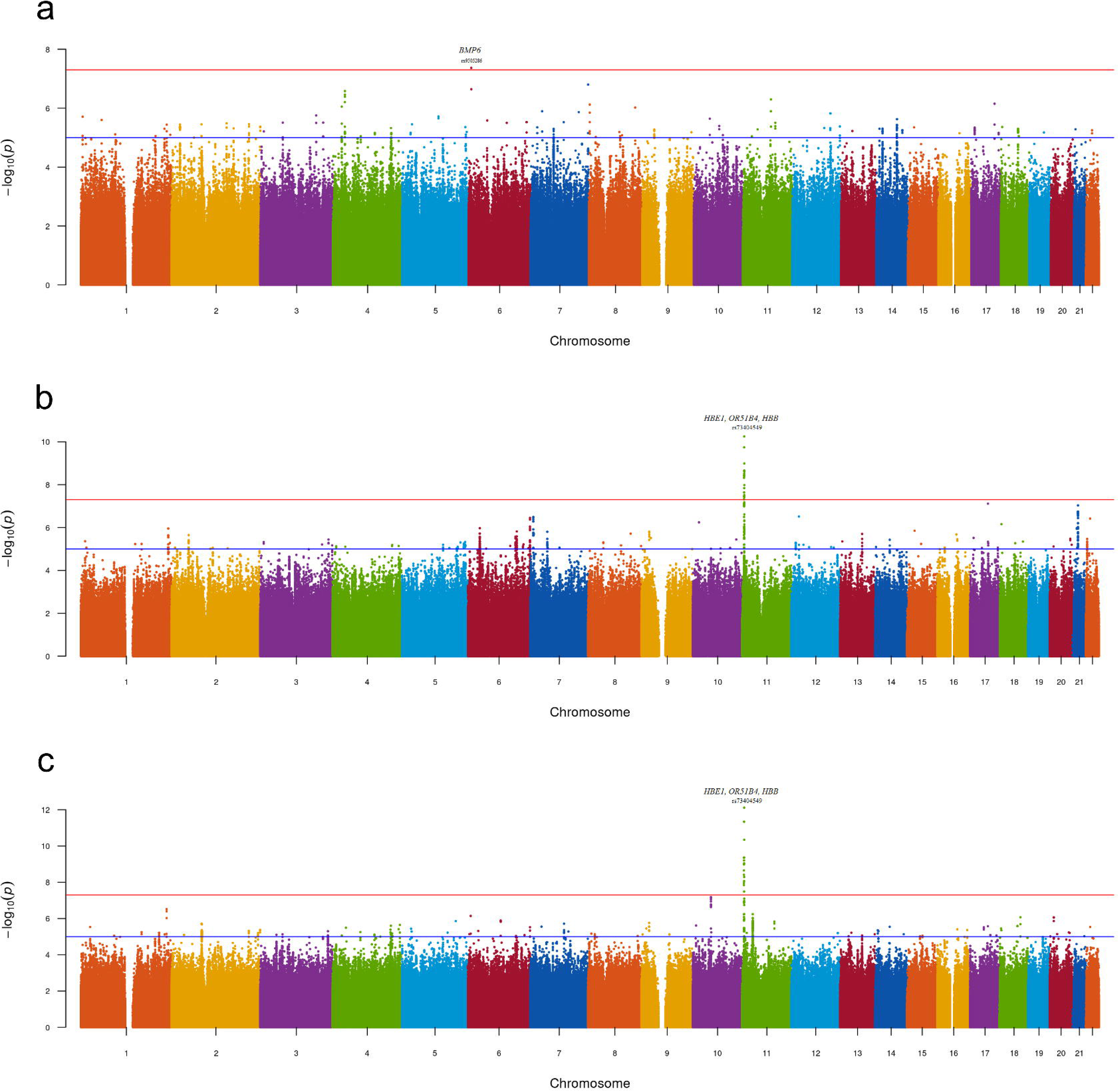
Manhattan plot - GWAS of UACR in the (a) Meta_SSA_ (b) Meta_NONRES_ (c) Meta_ALL_ datasets using the fixed effect model. Lead genome-wide significant SNPs (P<5×10^−08^) and gene annotations are highlighted.

No genomic inflation was observed for the individual-dataset association testing performed on the 9 datasets. All genomic inflation factors (lambda) were below 1.1. This was visually confirmed on the dataset specific QQ-plots and Manhattan plots (**Supplementary Figure S2 and Figure S3(a-i)**). Dataset-specific significant findings are reported in **Supplementary Table 2** and **Supplementary Figure S4(a-c)**.

One genome-wide significant locus with the lead SNP rs9505286 (p=4.3.10^−08^) was identified in Meta_SSA_ on chromosome 6. (**Figure 3a**). One genome wide significant locus with the lead SNP rs73404549 was identified on chromosome 11 in Meta_NONRES_ (p=5.6.10^−11^) and Meta_ALL_ (p=7.7 10^−13^) (**Table 2**, **Figure 3b and 3c**)

**Figure 3:**
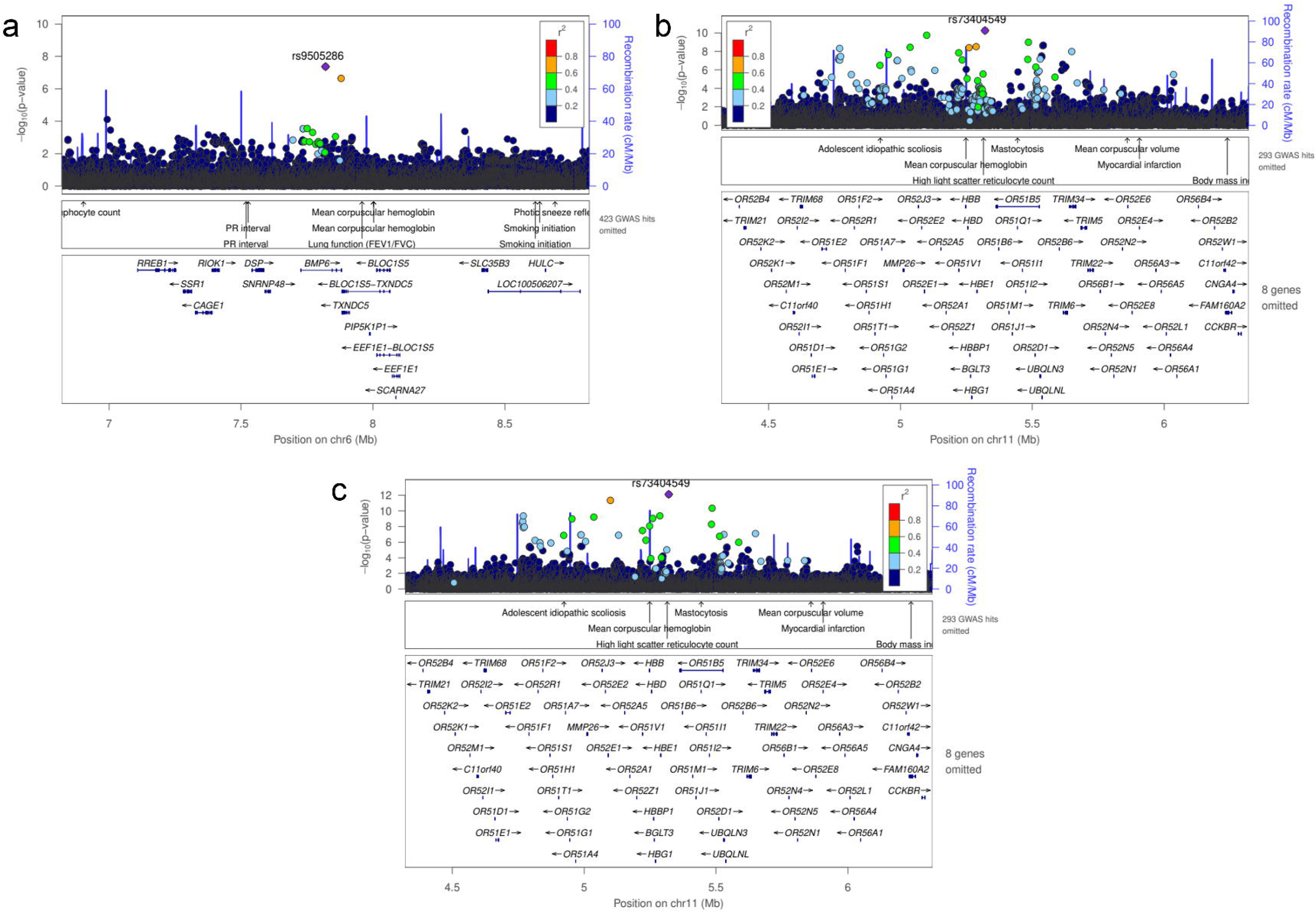
Regional plot using LocusZoom of genome-wide significant SNPs found in meta-analyses using the fixed effect model, (a) rs9505286 from the result of Meta_SSA_, (b) rs9966824 from the result Meta_NONRES_ (c) rs9966824 from the result Meta_ALL._

**Table 2:**
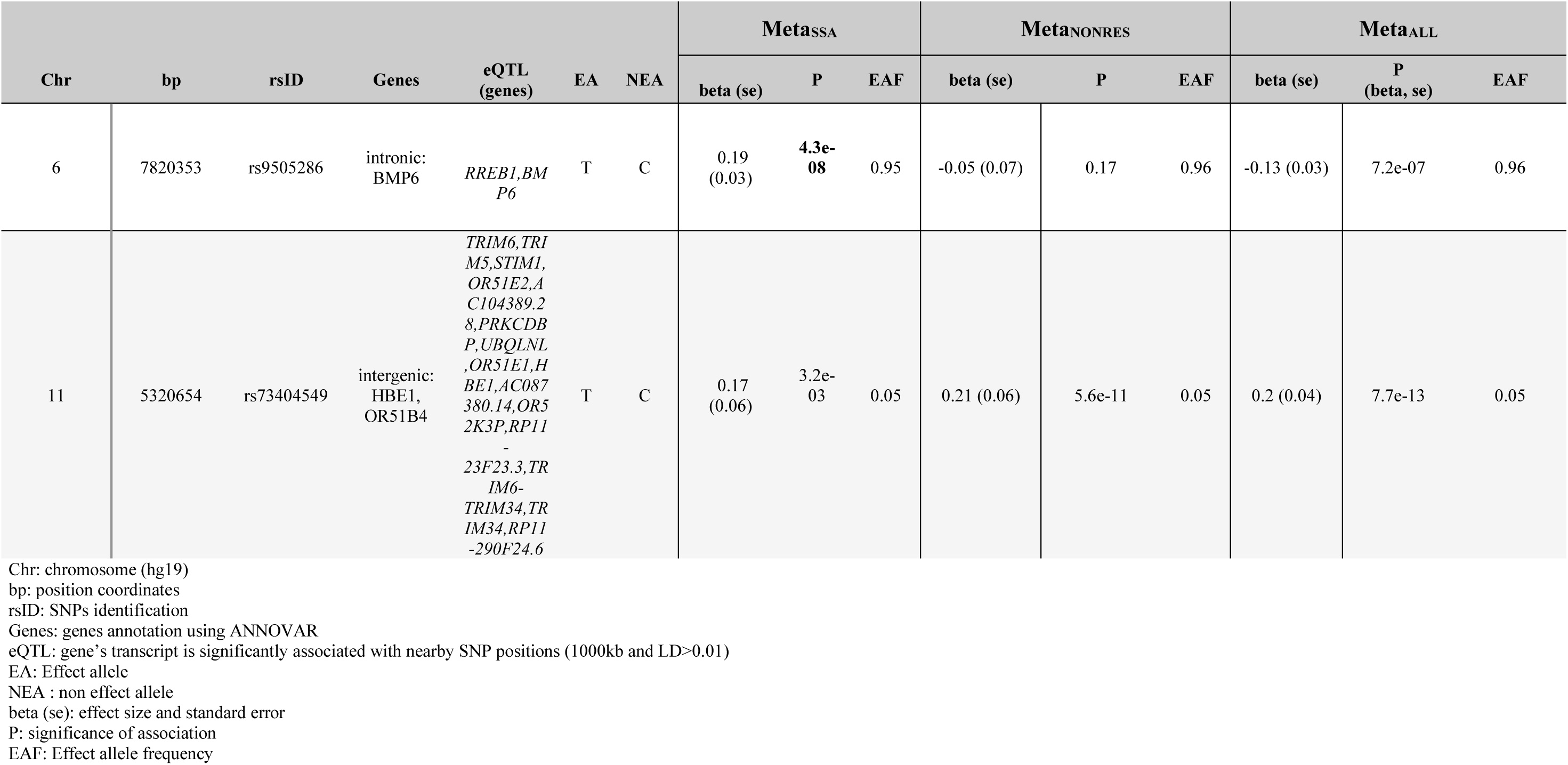
Lead genome-wide significantly associated SNPs for sub-Saharan African population meta-analysis (Meta_SSA_), non-resident African ancestry population meta-analysis (Meta_NONRES_) and the combined African ancestry population meta-analysis (Meta_ALL_).

SNP rs9505286 (chr6:7820353) is located in the intronic regions of *BMP6*. Two SNPs were identified in the 95% credible set using FINEMAP (**Figure 3a, Supplementary Table 4**). eQTLs in the region were found to be associated with the expression of two genes *RREB1* and *BMP6* (**Table 2; Supplementary Figure S5; Supplementary Table 3, Supplementary Table 5**).

SNP rs73404549 (chr11:5320654) is located near the *HBE1*, *OR51B4*, and *HBB* genes. This signal is primarily driven by results from West African ancestry datasets in the Meta_NONRES_ and Meta_ALL_ (**Figure 3b and 3c, Supplementary Figure S6**). Notably, this SNP is monomorphic in the Southern African and East African datasets. Furthermore, rs73404549 is in LD with rs334 (r^2^=0.52; 72,422 bp apart), the SNP that defines the sickle cell mutation (HbS). SNP rs334 was also significant in Meta_ALL_ (P_ALL_=8.55×10^−9^).

In the window of 1000kb around rs73404549, SNPs in the region colocalized with gene expression of *TRIM6* and *STIM1* in glomerular and tubulointerstitial tissues (**Table 2, Supplementary Table 5**).

Seven and two SNPs were identified in the 95 % credible set using FINEMAP in Meta_NONRES_ and Meta_ALL_ results respectively (**Supplementary Table 3; Supplementary Figure S6**).

### Replication of previous findings

Replication analysis confirmed associations in three previously identified regions in *THBS3*, *SPATA5L1/GATM*, and *ARL15* (**Supplementary Table 3**).

In the *THBS3* region, the Meta_ALL_ meta-analysis rs370545 was the lead SNP in our dataset, with a P-value of 1×10^−04^. However, a conditional analysis using rs2974937 (lead SNP in CKD-Gen-EA) resulted in a decrease in significance level (P_conditional_analysis_=0.85). This suggests that the association in the *THBS3* region was driven by rs2974937 in Meta_ALL_ even though it was not the lead SNP in this region (**Supplementary Table 4; Supplementary Figure 7**).

In the *ARL15* region, a statistically significant association signal was observed in Meta_SSA_ (rs1664781, P=1.8×10^−04^). Conditional analysis using rs1694068 (lead SNP in CDK-Gen-EA) revealed a reduction in P-value for rs1664781 (P_conditional_analysis_=0.87), suggesting that rs1694068 and rs1664781 are in LD thus confirming the association in this region (**Supplementary Table 4, Supplementary Figure 8**).

In the *SPATA5L1/GATM* region, the Meta_ALL_ meta-analysis identified rs1694067 as the lead SNP in this region with a P-value of 7.0×10^−05^. Furthermore, the lead SNP rs1153847 identified in CKD-Gen-EA, rs1153847, was present in our dataset, and its association was replicated (P_Bonferoni_adjusted_=0.04). For the window-based replication, a conditional analysis using rs2467858 (genome-wide significant SNP in CKD-Gen-EA), a reduced P-value was observed (P_conditional_analysis_=0.87) confirming rs1694067 and rs2467858 are in LD and replicated the CKD-Gen signal. (**Supplementary Table 4; Supplementary Figure 9**).

### Polygenic score analyses

The variance explained by the PGS for UACR residuals was between 0% and 0.82%. PGS constructed using the betas from CKD-Gen-EA and CKD-Gen-MA performed better for the non-SSA resident datasets, particularly in the UKB-African, showing the best predictivity (% variance: 0.82, p=1×10^−04^) and statistically significant correlation between the PGS and the UACR residual (**Figure 4, Supplementary Table S6**).

**Figure 4.**
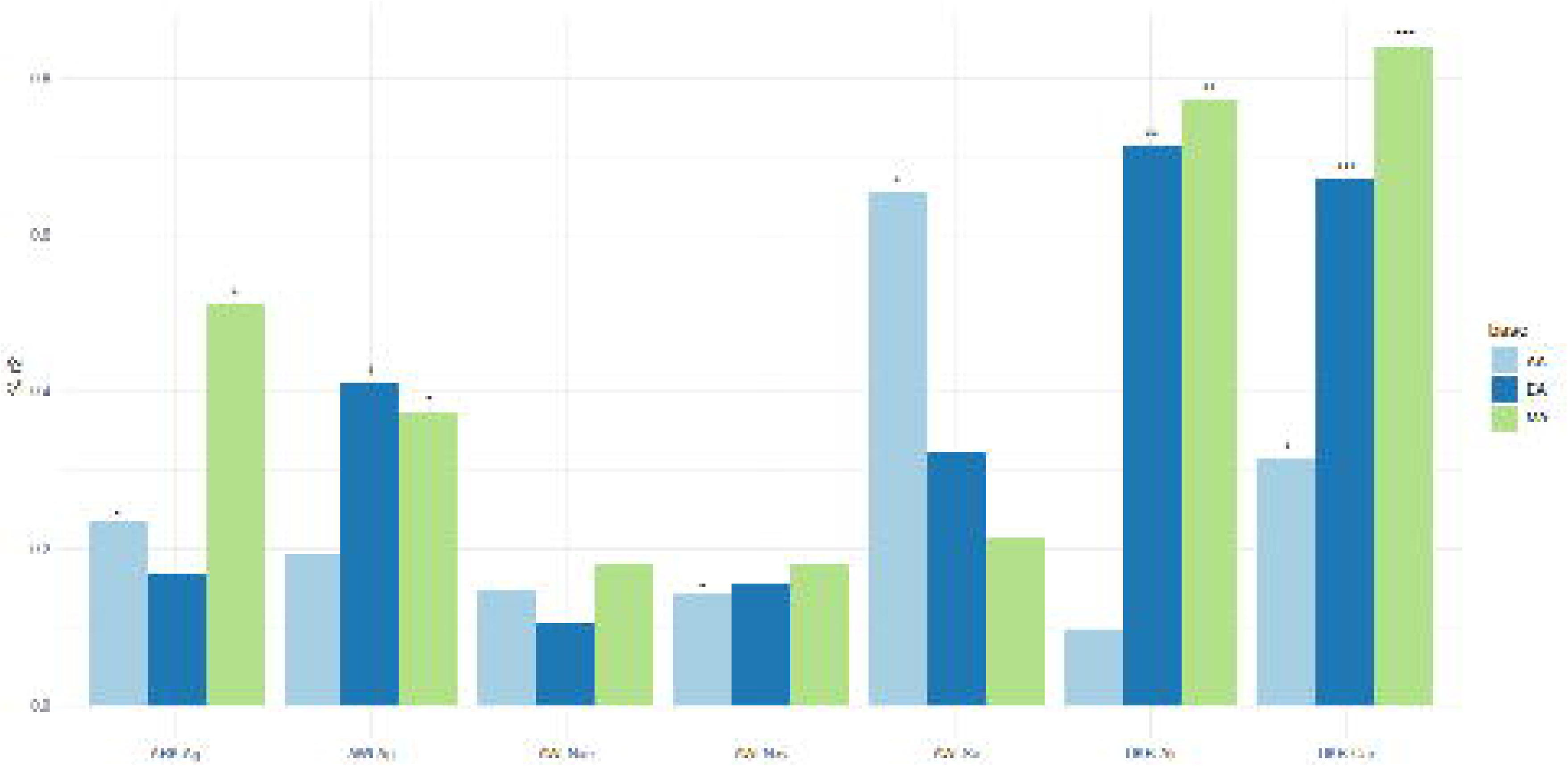
Percent variance explained between PGS and residual phenotypes computed using age, sex and 5 PCs. Key: The negative relationship between PGS and the phenotype in the result of the linear model, * P<0.05, ** P<0.01 and ** P<0.001. Details in Supplementary Table 6

Using the PGS constructed from CKD-Gen-MA, ARK-Agincourt (% variance: 0.61, P=0.01) and AWI-Agincourt (%variance: 0.58, P=0.002) demonstrated better predictivity in SSA populations. PGS constructed from CKD-Gen-AA did not improve the variance explained. Variance explained was lower using PGS constructed from CKD-GEN-AA than CKD-Gen-MA or CKD-Gen-EA.

## Discussion

This study is the first GWAS for UACR conducted in Sub-Saharan African populations. Two genomic regions were identified to be significantly associated with UACR among 8,970 participants from West, East, and Southern Africa and among 9,705 non-resident African-ancestry participants from the UK Biobank and CKD-Gen Consortium.

For the first locus, the SNP rs9505286, reached genome-wide significance in resident African individuals (MetaSSA) and is located in the intronic region of *BMP6*. eQTLs in LD with rs9505286 were found to be associated with expression of two genes, namely bone morphogenetic protein 6 (*BMP6*) and ras-responsive element binding protein 1 (*RREB1*). Both genes are plausibly linked with kidney disease. *BMP6* encodes a secreted ligand of the transforming growth factor (TGF-beta) superfamily of proteins, of which TGF-B1 is one of the most important regulators of kidney fibrosis, the pathological hallmark of irreversible loss of kidney function in CKD (Dendooven et al., 2011; Jenkins and Fraser, 2011). TGF-B1 is highly expressed in various fibrotic kidney diseases, including diabetic nephropathy (DN), hypertensive nephropathy, obstructive kidney disease, autosomal dominant polycystic kidney disease, immunoglobulin A nephropathy, crescentic glomerulonephritis, and focal segmental glomerulosclerosis. Because of its pivotal role in mediating kidney fibrosis, TGF-B1 is a potential target for drug discovery, and these results point towards similar potential in African populations for further exploration. *RREB1*, initially identified as a repressor of the angiotensinogen gene, is associated with type 2 diabetes in African Americans with end stage kidney disease(Bonomo et al., 2014). *RREB1* polymorphisms have been shown to interact with *APOL1*, and are implicated in fat distribution and fasting glucose, a potential explanation for the association with type 2 diabetes. As obesity and type 2 diabetes prevalence emerge in many African communities undergoing rapid sociodemographic transition, these findings must inform future work (Bonomo et al., 2014). Unfortunately, neither of the eQTLs has strong linkage disequilibrium (LD) support with the lead SNP rs9505286 (see Supplementary Table 5).

Variability of kidney function, confounding factors and allele frequency differences between datasets may explain why the rs9505286 signal was not replicated in Meta_ALL_ or Meta_NONRES_ (Marigorta et al., 2018).

For the second locus, the SNP rs73404549 was found to be statistically significant in non-resident individuals with African Ancestry (Meta_NONRES_) and overall (Meta_ALL_), but not in Sub-Saharan African individuals (Meta_SSA_). This can be explained by the fact that the variant allele of rs73404549 is extremely rare or absent in East and South African populations. This SNP was found to be in linkage disequilibrium with rs334, the sickle cell trait (HbS) in the *HBB* gene. The HbS mutation has been linked to malaria resistance among heterozygotes, with differences in allele frequency attributed to variations in selection pressures between Bantu-speaking populations in West and South/East Africa (Gurdasani et al., 2019; Choudhury et al., 2020). Notably, sickle cell trait and rs334 had been associated with various kidney function (eGFR) and kidney disease traits, including albuminuria, and chronic and end-stage kidney disease in African, African American and US Hispanic/Latino populations (Naik et al., 2014; Gurdasani et al., 2019; Fatumo et al., 2020; Masimango et al., 2022). Furthermore, an interaction between *APOL1* high-risk genotypes and the sickle cell trait enhances the risk for low eGFR(Masimango et al., 2022).

In addition to the *HBB* region, our GWAS revealed transferability of three previously identified signals. Of the 60 UACR-associated loci identified in European and Multi-Ancestry studies, only three were replicated, including variants in *GATM.* This region was also associated with eGFR in a Ugandan population (Fatumo et al., 2020). We also replicated the association with *ARL15* in the region of chromosome 1. *ARL15* is a regulator of Mg2+ transport thereby promoting the complex N-glycosylation of cyclin M proteins (CNNM 1-4) and could play a role in the pathogenesis of hypertension mediated via altered tubular handling of magnesium in the kidney (Zolotarov et al., 2021).

Allelic heterogeneity is high in African ancestry populations, as demonstrated by the high genetic diversity in our study (**Supplementary Fig. 1**). However, analysis of regional subgroups using meta-analysis (residents of South, West, or East Africa) did not reveal significant population-specific signatures (p<5e-8), likely due to small sample sizes within these subgroups (**Supplementary Fig. 10a, 10b, 3i**). Interestingly, meta-analysis under a random-effects model that allows for heterogeneity in allelic effects between regions (Meta_ALL_^RE^) did not improve the detection of specific signals already observed with the fixed-effects methods for HBB (Supplementary Figure S11). Consequently, the heterogeneity observed might be explained primarily by variations in linkage disequilibrium or environmental factors rather than by the effect of a specific allele, such as the presence or absence of sickle cell trait (Adriani Nikolakopoulou et al., 2014; Kuchenbaecker et al., 2019; Choudhury et al., 2020).

The transferability of PGS developed using the effect sizes quantified in three previous association studies in European ancestry, African ancestry and multi-ancestry populations showed limited predictability, explaining less than 1% of the variability in UACR. PGS in resident African populations (AWI-Gen and ARK) explained between 0.58% and 0.60% of the variance of UACR compared to UKB-African, where best prediction was observed (0.80%). The poor predictability of UACR using summary statistics derived from African Americans was likely due to the small sample size of the discovery dataset. Unfortunately, there have been few studies on PGS approaches to compare findings with, and the genetic heritability of UACR is relatively low, estimated at 4.3% (Teumer et al., 2019).

The limited transferability of PGS and previous GWAS signals across ancestral groups could be due to differences in genetic architecture and/or pleiotropic effects. Different demographic histories and genetic selection pressures between European and African populations could modify the ability to replicate previous GWAS results due to differences in allele frequencies between non-African and African populations, with generally lower LDs in African genomes. Environmental factors and variability in the prevalence and aetiology of kidney- and disease-related risk factors such as diabetes and hypertension (Fatumo et al., 2020) could also influence the genetic architecture of kidney disease in Africans populations (Limou et al., 2014; Teumer et al., 2019; Brandenburg et al., 2022b). Selection pressures have increased the frequencies *APOL1* kidney risk variants and HbS due to their protective properties in areas of Africa where trypanosomiasis and malaria are endemic. This may have contributed to shaping genetic susceptibility to kidney disease in African individuals. In our study, the *APOL1* gene region did not exhibit significant associations with UACR. The indel rs71785313 was not imputed using the African Sanger reference for imputation, and a specific study had previously been published to describe *APOL1* variant distribution in the AWI-Gen dataset using other imputation panels, but the locus did not reach genomic significance (5e^−8^) for association with eGFR and UACR (Brandenburg et al., 2022b).

While the burden of CKD in SSA is high, it is noteworthy that no prior GWAS on UACR has been conducted on the continent. Despite its uniqueness, our study is limited by its relatively modest sample size, which impacts statistical power to detect small-effect associations reaching genome-wide significance thresholds. Kidney and disease markers were measured at a single time point, and spot urine albumin and creatine levels are sensitive to incident infections and other environmental factors that could affect the prevalence of albuminuria.

It is important to note that our study populations are mainly treatment naïve in relation to kidney disease and other cardiometabolic conditions, which may be an advantage in detecting genetic associations (Pereira et al., 2021). Other studies, based on lipid-associated loci, attributed non-transferability of associated loci to pleiotropic effects, gene-environment interactions, and also to variability in allele frequencies and LD patterns (Kuchenbaecker et al., 2019; Choudhury et al., 2022), as we hypothesize for UACR.

In conclusion, this study describes genetic associations with UACR in a unique SSA cohort and non-resident individuals with African ancestry. CKD in African populations remains understudied but from available data, hypertension, rather than diabetes is the most commonly associated risk factor and in some regions, up to 60% of people with CKD do not have an associated “traditional” risk factor common to high-income settings, suggesting alternate underlying molecular pathways or aetiologies for CKD (Kalyesubula et al., 2018; Nakanga et al., 2019; Muiru et al., 2020). Our study identified two novel SNPs associated with UACR in populations of African ancestry. We further replicated three known UACR-associated loci. Regional genetic diversity due to different selection pressures appear to play a role in the genetic aetiology of CKD across the African continent. These factors likely contribute to the limited transferability of previous association signals and the poor transfer of polygenic scores developed in non-African populations to African populations. Larger genomic studies are necessary to better understand the genetic architecture of kidney function and chronic kidney disease across different African populations and inform region-specific kidney risk profiles. As demonstrated in this study, the low genetic heritability of UACR limits the predictive power of genetic risk scores (GRS) for kidney disease in our setting. It is critical for future research to address these gaps by modelling integrative risk scores that incorporate locally relevant clinical risk factors that are powerful predictors of kidney disease, multiple kidney phenotypes (eGFR_cystatin C_, eGFR_creatinine_, eGFR_creatinine + cystatin C_, albuminuria, Blood urea nitrogen), using multi-omics (Eddy et al., 2020), and the impacts of African-specific genetic risk for kidney disease, such as *APOL1* high-risk genotypes and sickle cell trait or disease(Naik et al., 2014; Friedman and Pollak, 2016; Brandenburg et al., 2022b).

## Ethics Statement

This study received approval from the Human Research Ethics Committee (Medical), University of the Witwatersrand/South Africa (M121029; M170880; M160939), the approval of the Centre Muraz Institutional Ethics Committee/Burkina Faso (015-2014/CE-CM) and the approval of the National Ethics Committee For Health Research/Burkina Faso (2014-08-096), the Ghana Health Service Ethics Review Committee (ID No: GHS-ERC:05/05/2015), the AMREF Health Ethics and Scientific Review Committee in Kenya (approval no. P114/2014) and the Navrongo Institutional Review Board (ID No: NHRCIRB178). All the participants signed an Informed Consent Form before any study procedure was performed.

## Acknowledgement

We acknowledge the sterling contributions of our field workers, phlebotomists, laboratory scientists, administrators, data personnel, and all other staff who contributed to the AWI-Gen and ARK data and sample collection, processing, storage, and shipping, and the participants who generously agreed to be part of the studies. The UK Biobank kindly shared data under the UK Biobank Resource Application Number 63215 and approved submission of the paper.

## Author Contributions

JTB and MR designed the study, JTB performed the analyses and wrote the first draft. WCC, MR, AM, SH and JF made significant contributions to guiding the analyses and interpreting the results, as well as writing and reviewing the manuscript. All authors read, edited and approved the manuscript.

## Data availability statement

Summary statistics are available in the GWAS catalog (XXX) and scripts are available from the corresponding authors. The AWI-Gen data set is available from the European Genome-phenome Archive (EGA) database (https://ega-archive.org/), accession number EGAS00001002482 (phenotype dataset: EGAD00001006425; genotype dataset: EGAD00010001996). The availability of these datasets is subject to controlled access through the Data and Biospecimen Access Committee of the H3Africa Consortium. ARK data is available in WIReDSpace repository on request (https://wiredspace.wits.ac.za/), https://doi.org/10.54223/uniwitwatersrand-10539-3301635

Data are available under the terms of the Creative Commons Zero “No rights reserved” data waiver (CC0 1.0 Public domain dedication). The processed data generated in this study are provided in Supplementary Material. Permission was obtained to access the genotype and phenotype dataset for UKBB (research project number: 63215).

Publicly available databases include Teumer et al. in CKD-GEN consortium website: https://ckdgen.imbi.uni-freiburg.de/.

