## Supplementary Figure for "Genetic Association and Transferability for Urinary Albumin-Creatinine Ratio as a Marker of Kidney Disease in four Sub-Saharan African Populations and non-continental Individuals of African Ancestry"

Jean-Tristan Brandenburg

April 4, 2024

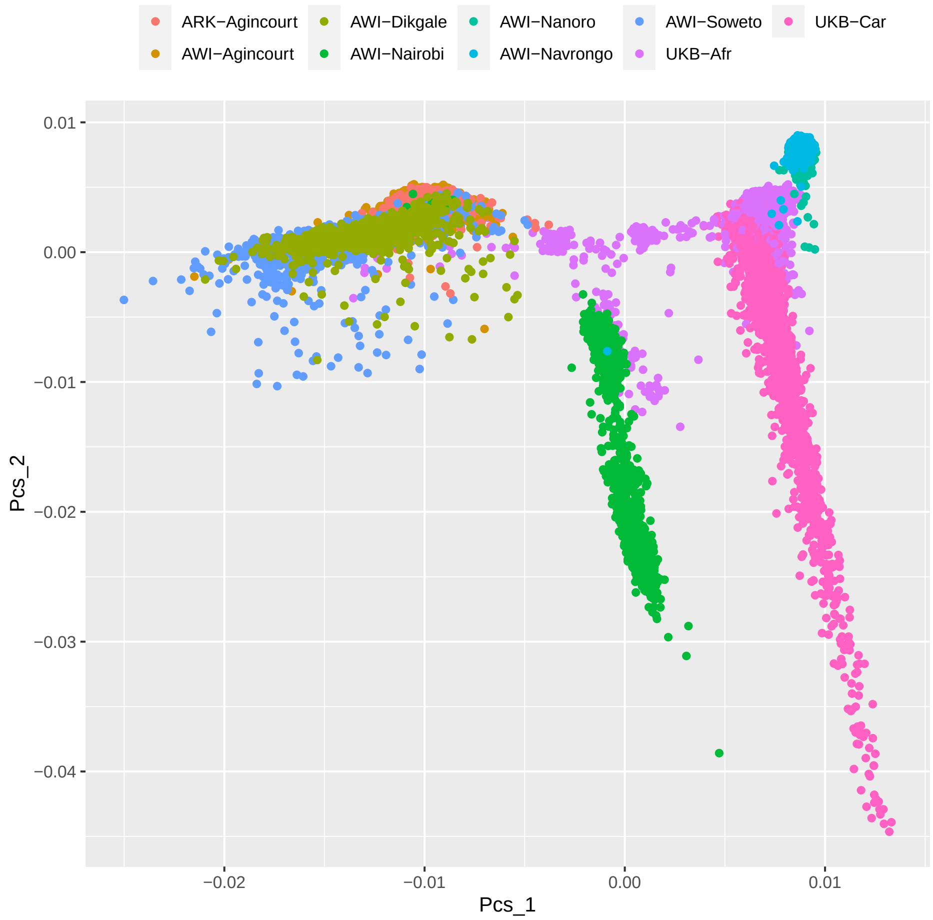

Figure S1: Genetic Diversity of pooled samples using PC1 and 2

AWIGEN: Africa Wits-INDEPTH partnership for Genomic Studies ; ARK: African Research on Kidney Disease Study;UKB: UK Biobank

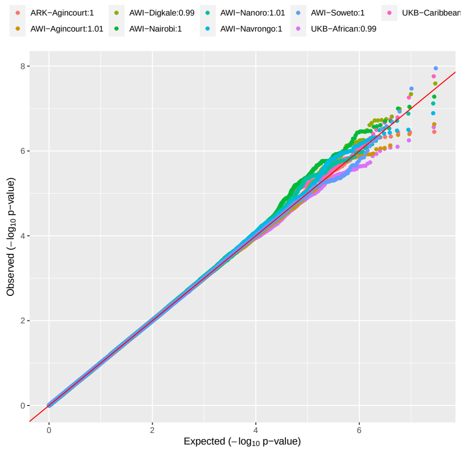

Figure S2a: QQ plot and lambda value for UKB-African, UKB-Caribbean, AWI-Agincourt, ARK-Agincourt, AWI-Dikgale, AWI-Navrongo, AWI-Nanoro, AWI-Soweto and AWI-Nairobi

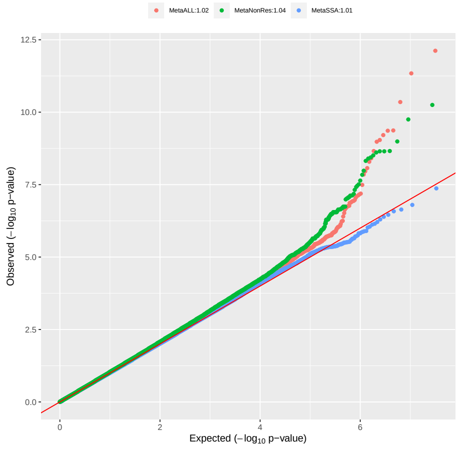

Figure S2b: QQ plot and lambda value for $\mathrm{Meta}_{\mathrm{SSA}}$, $\mathrm{Meta}_{\mathrm{ALL}}$ and $\mathrm{Meta}_{\mathrm{NONRES}}$

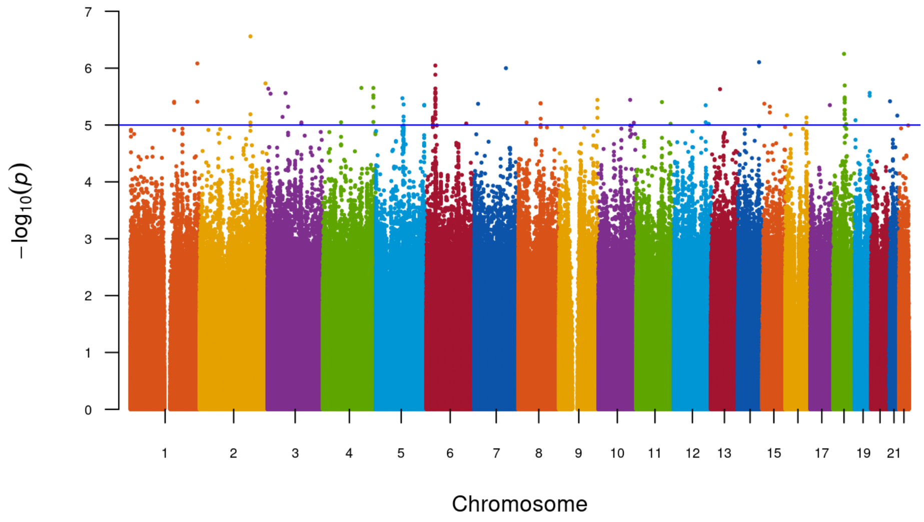

Figure S3a: Manhattan plot for UKB-African

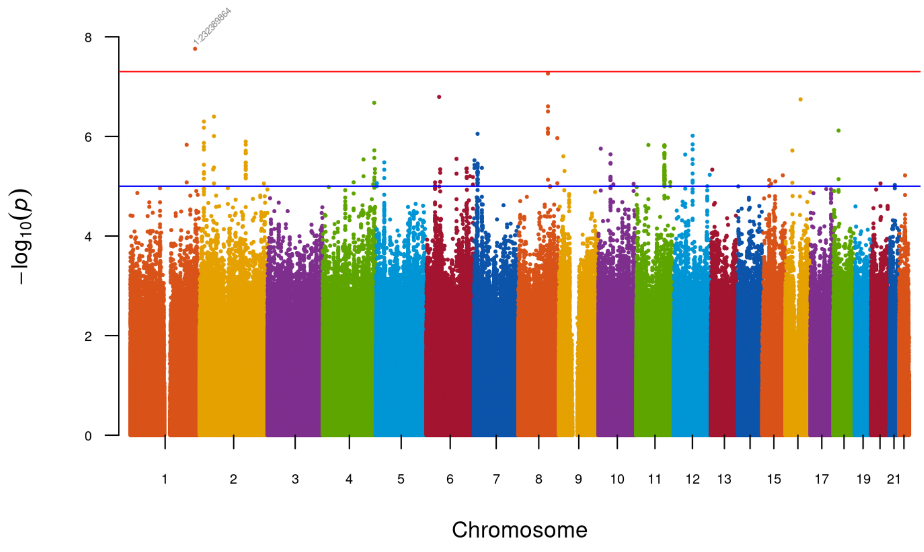

Figure S3b: Manhattan plot for UKB-Caribbean

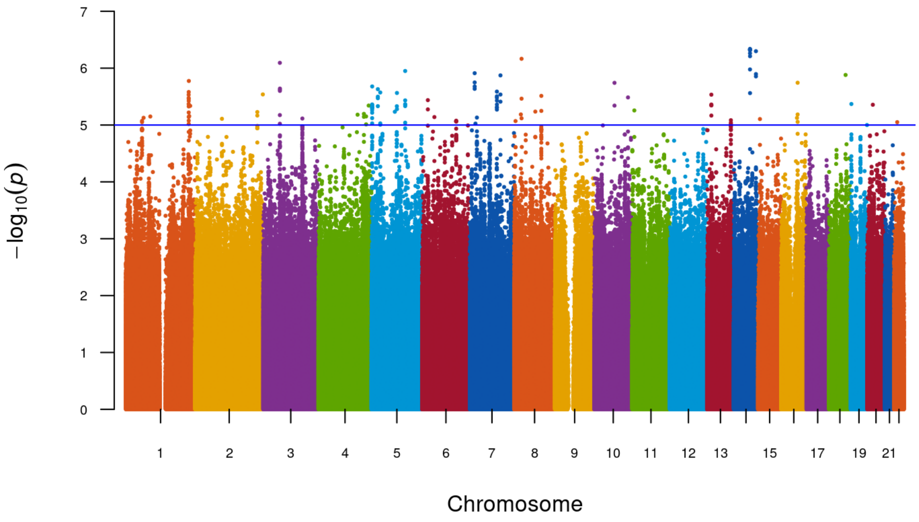

Figure S3c: Manhattan plot for ARK-Agincourt

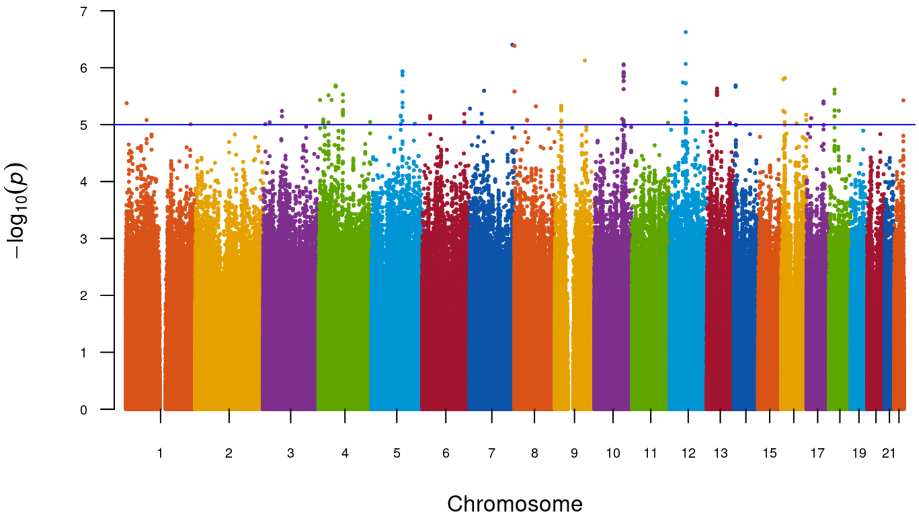

Figure S3d: Manhattan plot for AWI-Agincourt

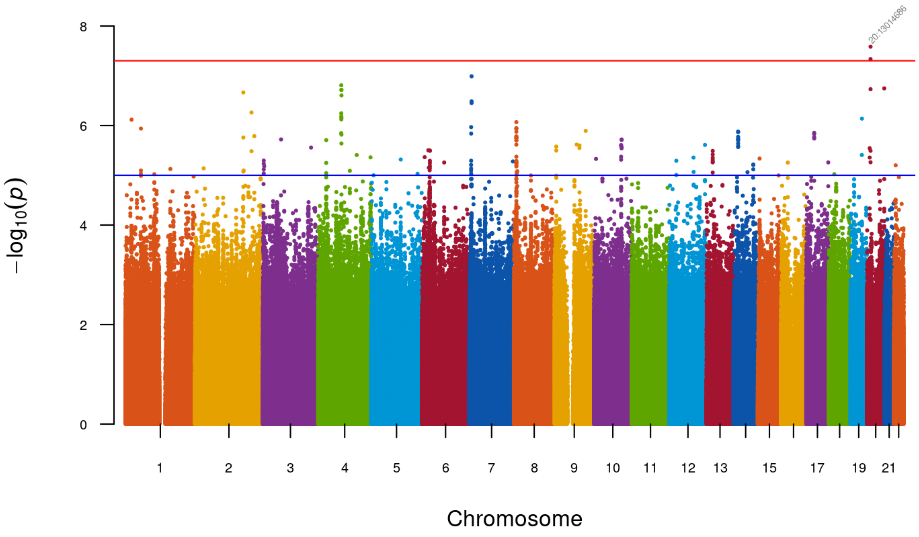

Figure S3e: Manhattan plot for AWI-Dikgale

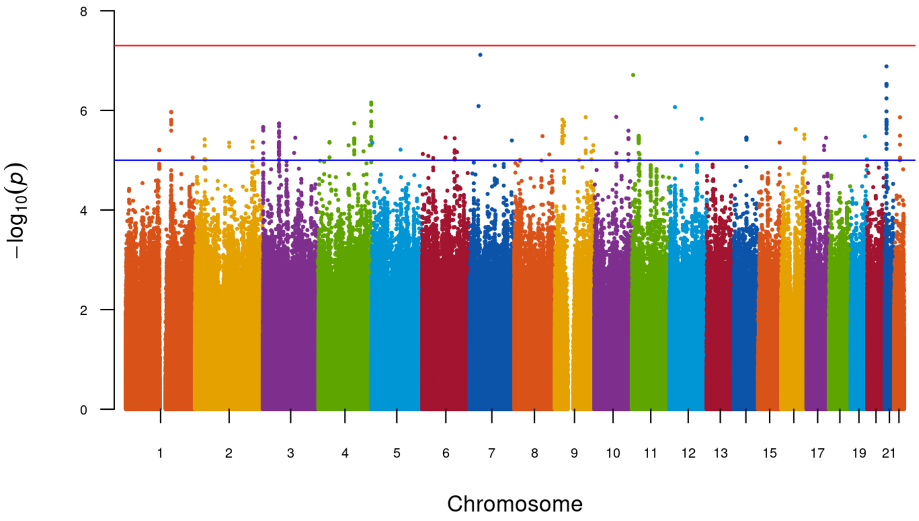

Figure S3f: Manhattan plot for AWI-Nanoro

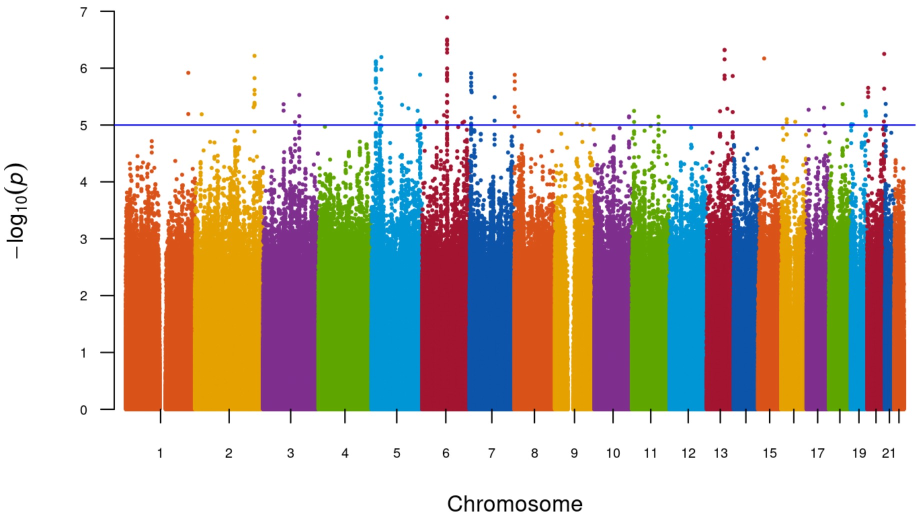

Figure S3g: Manhattan plot for AWI-Navrongo

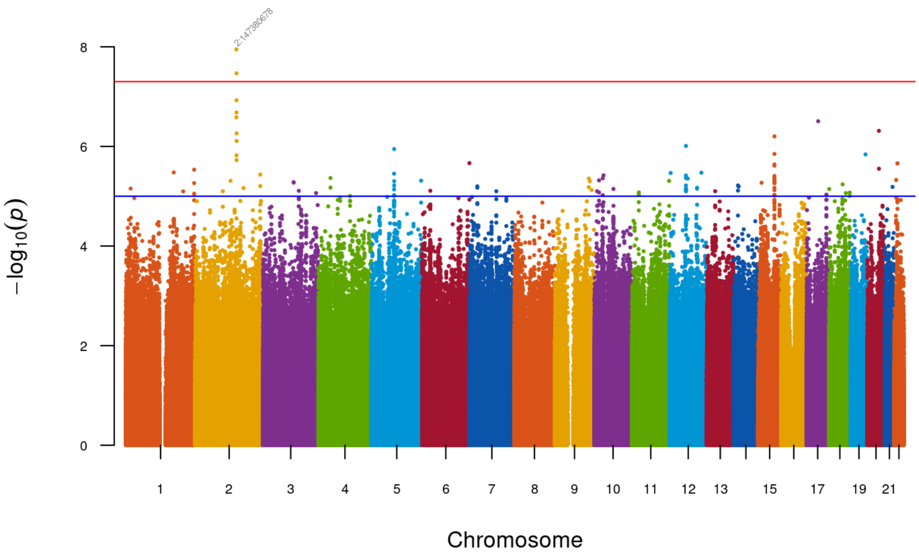

Figure S3h: Manhattan plot for AWI-Soweto

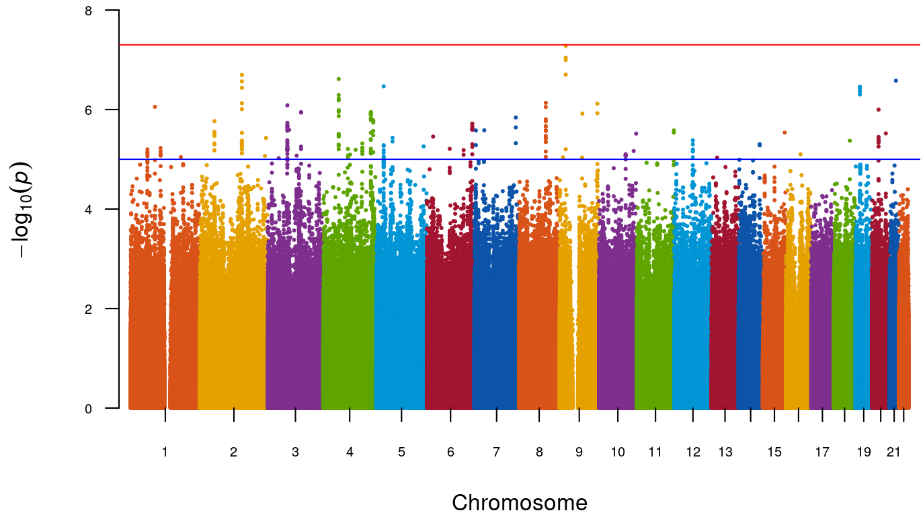

Figure S3i: Manhattan plot for AWI-Nairobi

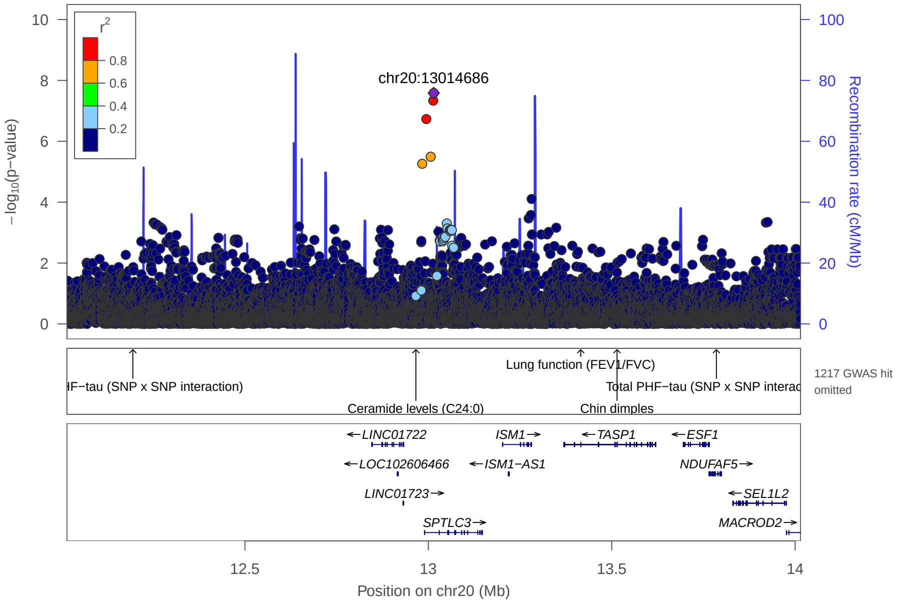

Figure S4a: Regional plot around rs2052976 (20-13014686) using AWI-Dikgale results
rs2052976 found significant in AWI-Dikgale, regional plot had been done using locuszoom with a windows of 1MB around and LD had been estimated using sample from AWI-Dikgale

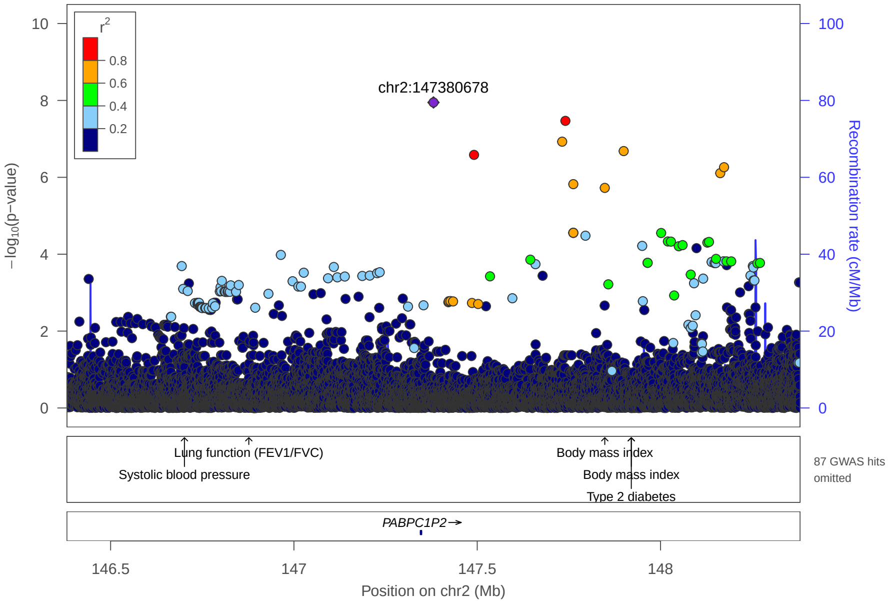

Figure S4b: Regional plot around rs147938214 (2-147380678) using AWI-Soweto results

rs147938214 found significant in AWI-Soweto, regional plot had been done using locuszoom with a windows of 1MB around and LD had been estimated using sample from AWI-Soweto

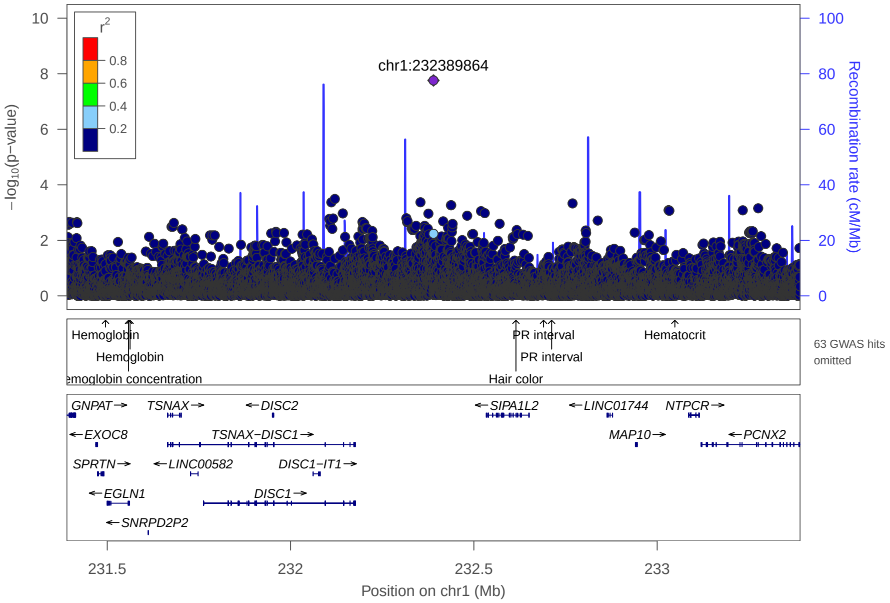

Figure S4c: Regional plot around rs12067862 (1-232,389,864) using UKB-Caribbean results

rs12067862 found significant in UKB-Caribbean, regional plot had been done using locuszoom with a windows of 1MB around and LD had been estimated using sample from UKB-Caribbean

| Study | af | P | N |
|---|---|---|---|
| Ark−Agincourt | 0.945 | 0.13 | 928 |
| AWI−Agincourt | 0.951 | 3.4e−03 | 1545 |
| AWI−Soweto | 0.965 | 0.17 | 766 |
| AWI−Dikgale | 0.967 | 0.25 | 917 |
| AWI−Nanoro | 0.943 | 0.01 | 1702 |
| AWI−Navrongo | 0.961 | 7.0e−03 | 1548 |
| AWI−Nairobi | 0.937 | 0.06 | 1481 |
| **MetaSSA** | **0.951** | **4.3e−08** | **8887** |
| UKB−African | 0.949 | 0.1 | 1204 |
| UKB−Caribbean | 0.956 | 0.09 | 1710 |
| CKDGEN−AA | 0.966 | 0.11 | 6795 |
| **MetaNONRES** | **0.962** | **0.17** | **9709** |
| **MetaALL** | **0.957** | **7.2e−07** | **18596** |

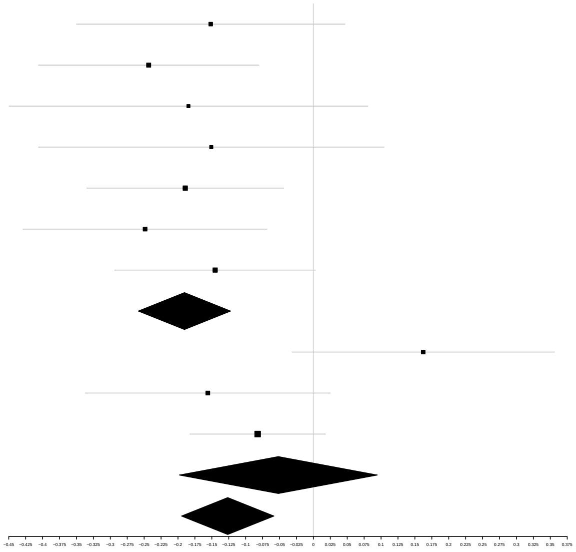

Figure S5: Forest plot (beta, se) and af, p-value and N for each dataset and meta analysis result of rs9505286 (chr6,7820353) found significant in $\text{Meta}_{\text{SSA}}$

| Study | af | P | N |
|---|---|---|---|
| Ark−Agincourt | | | |
| AWI−Agincourt | | | |
| AWI−Soweto | | | |
| AWI−Dikgale | | | |
| AWI−Nanoro | 0.04 | 0.13 | 1702 |
| AWI−Navrongo | 0.053 | 8.8e−03 | 1548 |
| AWI−Nairobi | | | |
| **MetaSSA** | **0.046** | **3.2e−03** | **3250** |
| UKB−African | 0.089 | 6.1e−05 | 1204 |
| UKB−Caribbean | 0.053 | 3.1e−05 | 1710 |
| CKDGEN−AA | 0.046 | 1.5e−04 | 6795 |
| **MetaNONRES** | **0.053** | **5.6e−11** | **9709** |
| **MetaALL** | **0.051** | **7.7e−13** | **12959** |

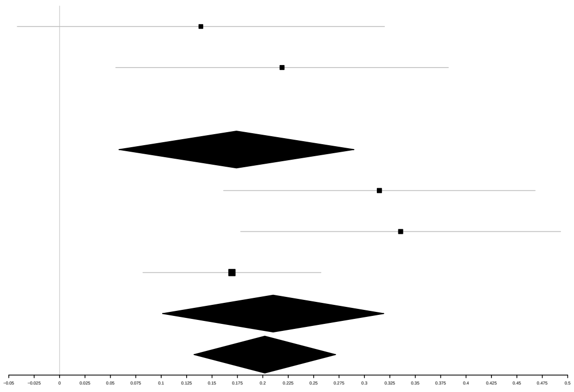

Figure S6: Forest plot (beta, se) and af, p-value and N for each dataset and meta analysis of rs73404549 (chr11,5320654) found significant in $\mathrm{Meta}_{\mathrm{NONRES}}$, $\mathrm{Meta}_{\mathrm{ALL}}$

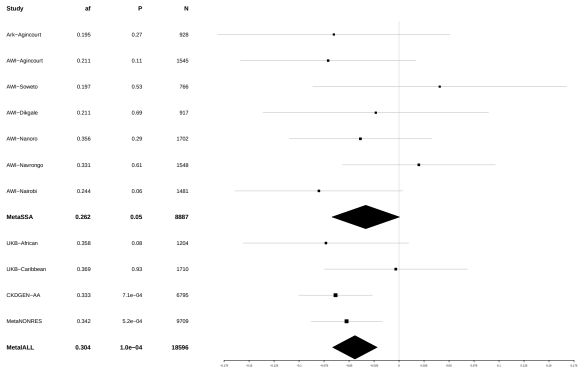

(a) Forest plot of rs370545

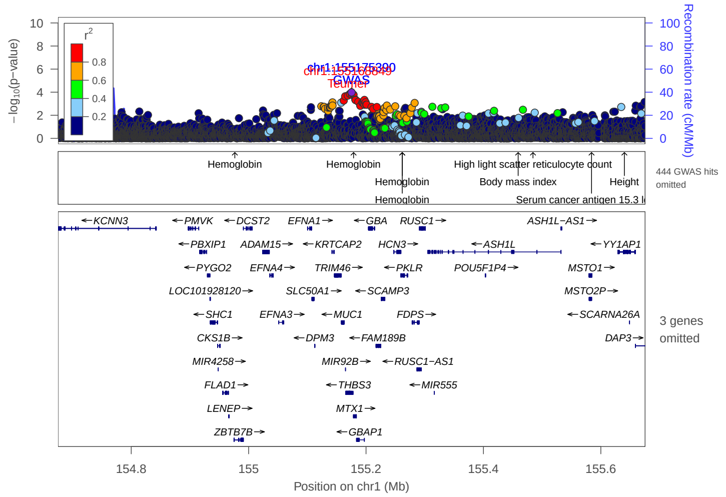

(b) Regional plot around rs370545 in $\text{Meta}_{\text{ALL}}$

Figure S7: Replication of *THBS3* regions identified in CKDGEN-EA : forest plot of lead SNPs identified in $\text{Meta}_{\text{SSA}}$ rs370545 (a) and regional plot around (b)
regional plot had been perfomed using locus zoom, using $\text{LD}_{\text{ALL}}$, lead SNPs identified in $\text{Meta}_{\text{ALL}}$ are annotated in blue and Significant SNPs identified in CKDGEN-EA are highligth in red (b)

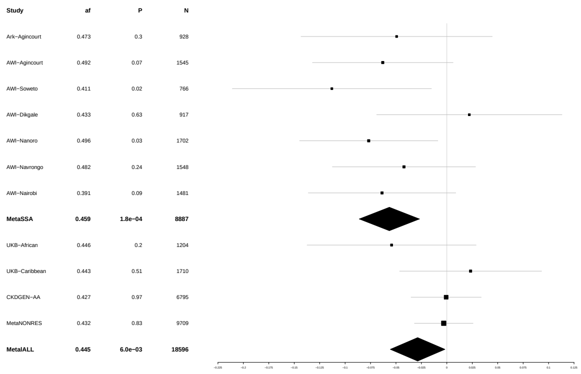

(a) Forest plot of rs1694067

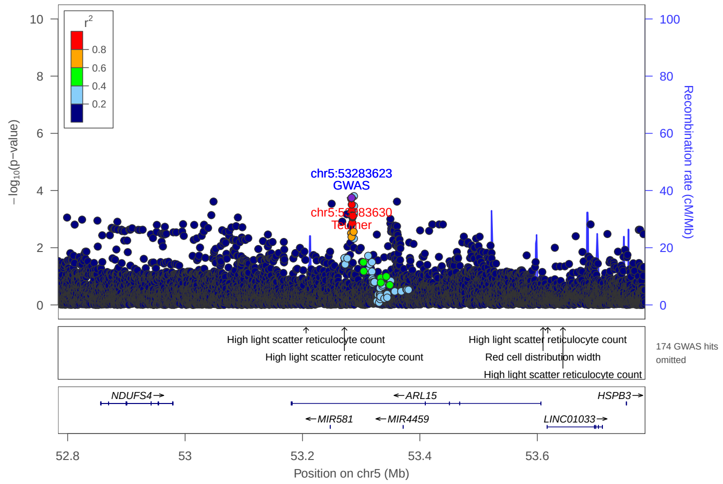

(b) Regional plot using Meta$_{SSA}$ and CKDGEN-MA

Figure S8: Replication of *ARL15* regions identified in CKDGEN-EA and CKDGEN-MA : forest plot of lead SNPs identified in Meta$_{SSA}$ rs1694067 (a) and regional plot around (b)
regional plot had been perforned using locus zoom, using LD$_{SSA}$, lead SNPs identified in Meta$_{SSA}$ are annotated in blue and Significant SNPs identified in CKDGEN-MA are highligth in red (b)

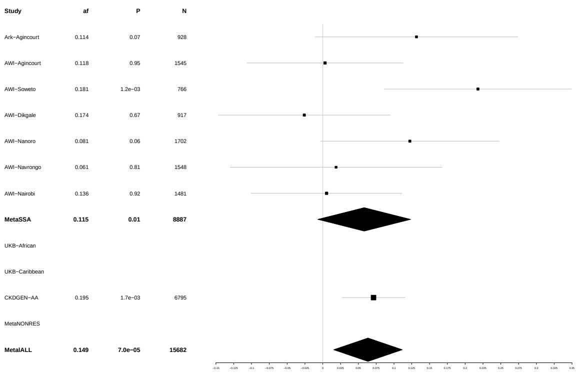

(a) Forest plot of rs1153847

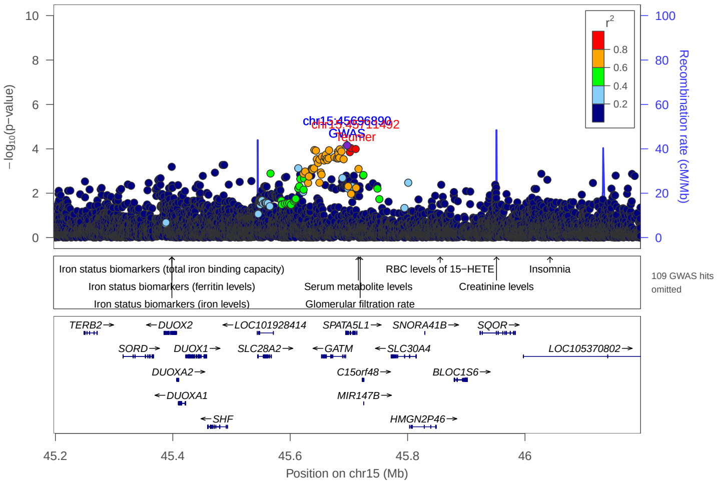

(b) Regional plot around rs1153847 using Meta$_{\mathrm{ALL}}$ and CKD-GEN-EA

Figure S9: replication of *GATM/SPATA5L1* regions identified in CKDGEN-EA : forest plot of lead SNPs identified in Meta$_{\mathrm{ALL}}$ rs1153847 (15-45696890) (a) and regional plot around (b)
regional plot had been perforned using locus zoom, using LD$_{\mathrm{ALL}}$, lead SNPs identified in Meta$_{\mathrm{ALL}}$ are annotated in blue and Significant SNPs identified in CKDGEN-EA are highligth in red (b)

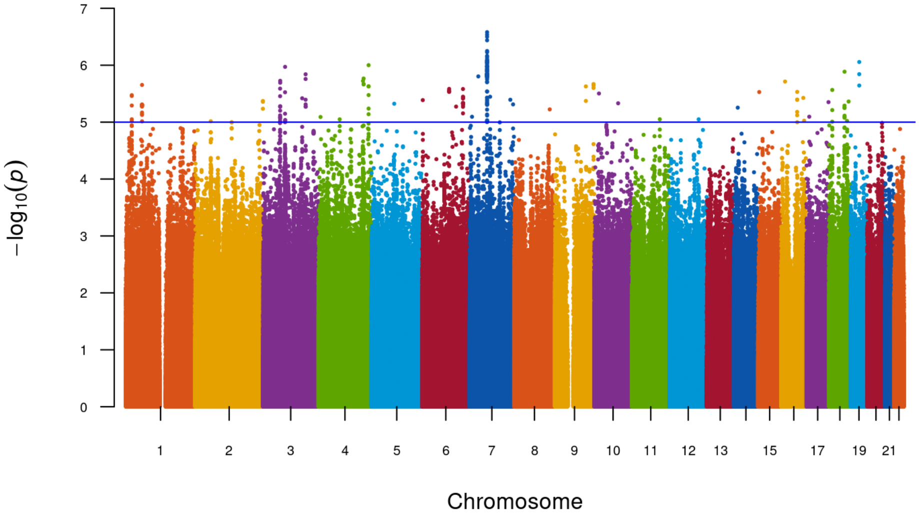

Figure S10a: Manhattan plot of Meta Analysis using South resident using Fixed effect method

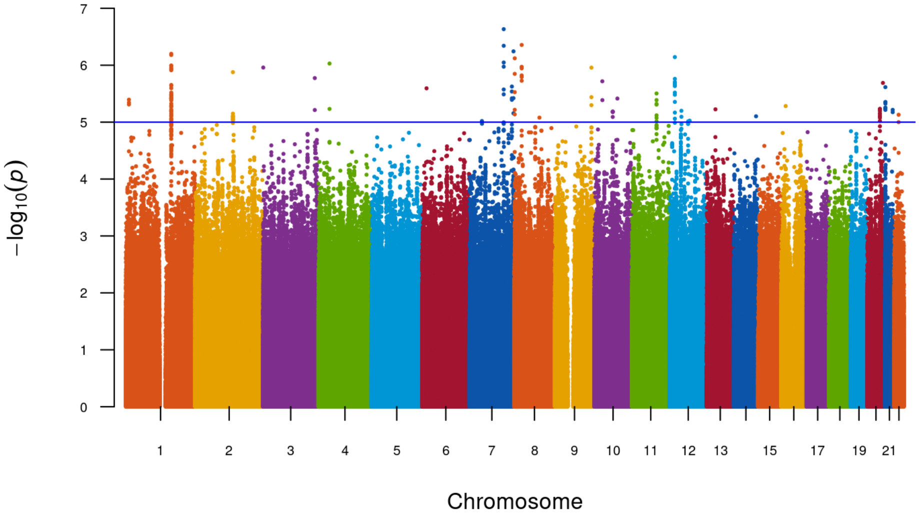

Figure S10b: Manhattan plot of Meta Analysis using West resident using Fixed effect method

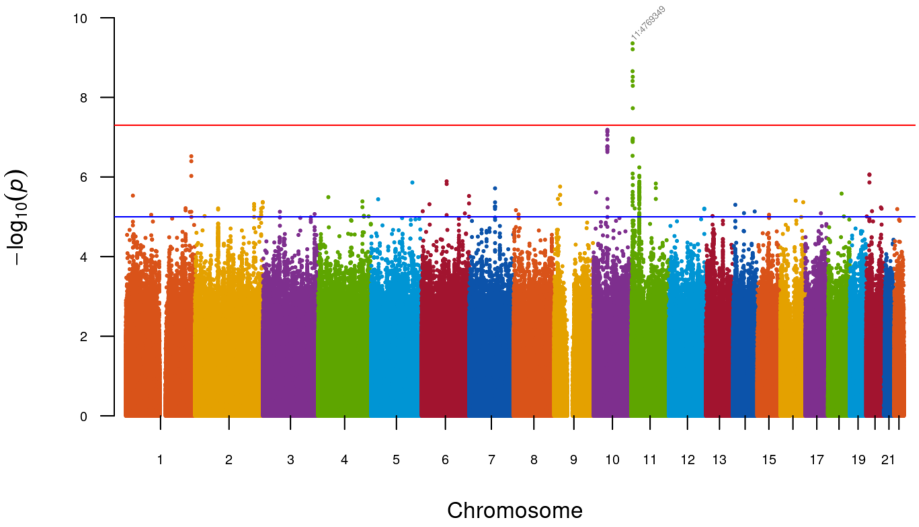

Figure S11: Manhattan plot of Meta Analysis using all dataset using Random effect
